# Analysis of Premature Termination Codons Predicted to Escape Nonsense Mediated Decay Identifies Novel Genes, Pathways, and Networks Contributing to an Oligogenic Etiology of Congenital Heart Disease

**DOI:** 10.1101/2025.08.04.25332980

**Authors:** Jonathan Klonowski, Madhavi Ganapathiraju, Archana Rai, Ben Glennon, Zeynep Coban-Akdemir, Dennis Kostka, Cecilia Lo

## Abstract

**Background:** A large fraction of clinically relevant pathogenic genomic variation consists of premature termination codons (**PTC).** While PTCs can trigger nonsense mediated mRNA decay (**NMDtrig**) causing loss of function (**LOF**), those near the end of transcripts can escape NMD (**NMDesc**), allowing expression of truncated proteins. As NMDesc PTCs are not well studied, and can lead to possible gain of function (**GOF**) effects, we investigated the impact of NMDtrig/NMDesc PTCs on human congenital heart disease (**CHD**).

**Results:** Whole exome sequencing data from ∼3,000 CHD patients and ∼5,000 control subjects were analyzed for PTCs using known rules for classifying variants as NMDtrig/NMDesc. CHD patients had an increased burden of PTCs in gene-sets related to both heart and brain developmental processes. NMDesc PTCs were enriched in the *MAPK* pathway, known to harbor GOF variants that cause Noonan syndrome. Also identified were Hedgehog and ERBB signaling, pathways with prominent roles in heart development but not previously reported in nonsyndromic CHD. NMD analysis of PTCs at the transcript level identified 11 genes (9 novel) associated with CHD. Enrichment of digenic combinations between these 11 genes and digenic and higher order combinations of PTCs within heart developmental genes suggest a role for oligogenicity.

**Conclusions:** We observed NMDesc PTCs play a role in CHD pathogenesis, and this role may be disproportionately specific to heart development. Our analysis uncovered new genes and pathways, identifying MAPK, Hedgehog, and ERBB signaling as pathways contributing to nonsyndromic CHD. The additional observation of digenic and higher order combinations of PTCs suggests an oligogenic framework to the genetic architecture of human CHD.

## Introduction

Congenital heart disease (CHD), a birth defect involving structural abnormalities in the heart and great vessels,^1^ is observed in approximately 1% of live births and is among one of the most common birth defects.^2, 3, 4, 5^ Despite over 140 genes having been linked to human CHD and many others identified through analysis of animal models, ∼56% of human CHD cases remain unexplained. Despite the increasing availability of sequencing data from large CHD cohorts, it is difficult to distinguish between benign variants and those that may be pathogenic. Adding to the complexity is CHD’s sporadic occurrence, variable expressivity and incomplete penetrance.^6^ As a result, the genetic landscape of CHD is still not well understood.

One productive avenue of investigation has been the study of *de novo* variants, which led to the identification of chromatin modifiers as a significant contributor to CHD pathogenesis.^7^ However, as *de novo* variants are estimated to account for only 10% of pathogenic variants mediating CHD,^8^ the study of both *de novo* and inherited pathogenic variation is necessary to fully explain the genetic landscape of CHD. While such studies are challenging, a common strategy has been to focus on high-impact variants, such as nonsense, splice site, and frameshift variants (a grouping referred to as protein truncating variants; PTVs).^9^ While PTVs within the same gene are largely presumed to act in a single direction and through loss of function (LOF) effects, this is not always the case.

Nonsense and, in most cases, frameshift variants lead to premature termination codons (PTC) and are censored by the evolutionarily conserved nonsense mediated decay (NMD) system, which selectively degrades transcripts with PTCs. This system is imperfect and PTCs near the end of a transcript are frequently observed to escape NMD (NMDesc) and lead to expression of altered protein. These altered proteins can have altered activity, yielding again-of-function (GoF) allele (Figure 1a). As ∼36% of human disease-associated PTCs are predicted NMDesc variants, this would suggest such PTCs may have specific clinical significance. Supporting this possibility is the fact that PTCs in the same gene can lead to different clinical diseases based on whether the PTCs is positioned such that it triggers or evades NMD detection (NMDtrig vs. NMDesc).^10–13^ As recent studies examining loss-of-function (LoF) variants causing CHD sometimes outright excludes PTCs predicted to cause NMDesc,^14,15^ the potential contribution of NMDesc PTCs in CHD on the genomic scale remains unexplored.

**Figure 1.**
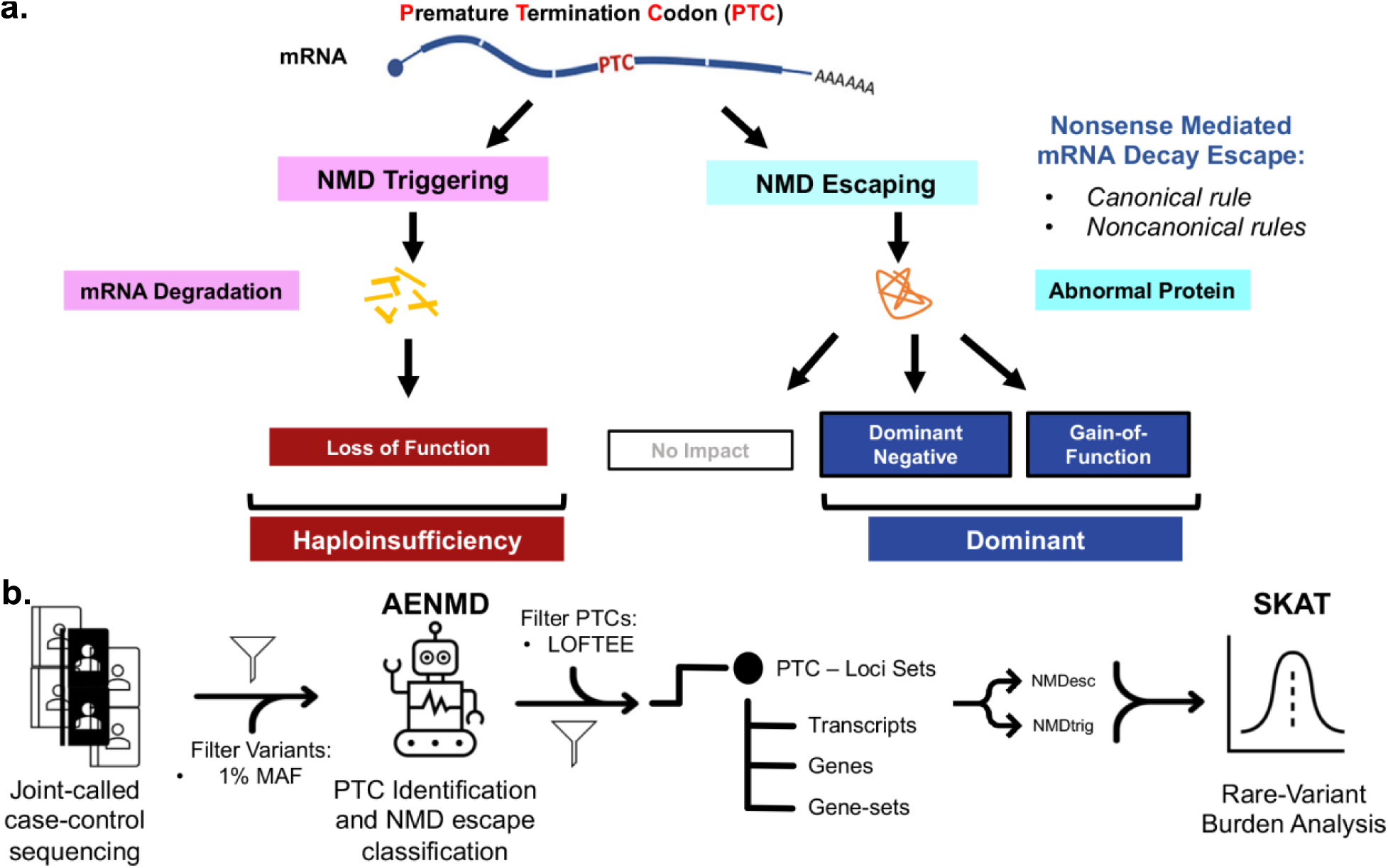
The biological and methodological processing of premature termination codons (PTC) (**a**) Illustrates the way PTCs are detected, or evade detection, by nonsense mediated mRNA decay (NMD) and potential molecular outcomes, often associated with dominant models of disease. (**b**) Illustrates the variant processing and analysis pipeline. Starting with joint-called case-control sequencing data and ending with rare-variant burden analysis to identify associations between variant sets and congenital heart disease (CHD)

At present, there is only a single well documented association of GOF genetic basis in CHD, whereby GOF pathogenic variants in the RAS/MAPK pathway have been identified to be associated with Noonan’s syndrome. Noonan’s syndrome is a syndromic disorder characterized by craniofacial anomalies, CHD (most often pulmonary stenosis), and other extracardiac anomalies.^16,17^ Herein we detail our systematic analysis of PTCs in CHD, investigating the individual contribution of NMDtrig and NMDesc PTCs. This analysis showed a burden of NMDesc PTCs in the RAS-MAPK pathway in CHD patients, supporting possible GOF effects of these variants. In addition, we recovered other cell signaling pathways not previously associated with human CHD. Gene-level analysis of NMDtrig and NMDesc PTCs recovered 9 novel CHD genes, and we observed an enrichment of CHD patients with digenic pairs and higher order oligogenic combinations of PTCs. These findings show efficacy in the analysis of NMDesc PTCs for interrogating the non-Mendelian genetic architecture of human CHD.

## Results

### Annotation of PTCs with AENMD in CHD Case-Control Cohort

Whole exome sequencing (WES) data from 3,081 CHD patients and 5,140 healthy subjects from the Alzheimer’s Disease Sequencing Project (ADSP) were joint called and analyzed in a case-control study design. The CHD cohort was composed of 3,081 subjects, 2,425 from the Pediatric Cardiac Genomics Consortium (PCGC) and 656 subjects recruited from Children’s Hospital of Pittsburgh (CHP). Common variants with an in-cohort or gnomAD minor allele frequency >1% were removed, leaving rare variants that were then processed by AENMD to recover PTC variants (Figure 1b). The PTC recovered were also classified on a per transcript basis by AENMD as either predicted as triggering NMD (NMDtrig) or evading NMD detection (NMDesc) using four previously experimentally validated rules for NMD based on transcript exon-exon structure.^13^ Together this delineates three NMDesc PTC groups: (1) coding start site proximal (CSS) PTCs; (2) PTC within exons spanning more than 407 base pairs (>407); and, (3) exon-junction complex dependent NMD escape region (EJC) PTCs. PTCs were further annotated using LOFTEE to flag and remove low quality PTCs without removing PTCs flagged for predicated NMD escape.^15^ This yielded 28,550 unique PTC-causing variants with a total allele count of 74,335 (Additional File 1: Table S1).

### Gene-set Enrichment of Variants in Heart and Brain Developmental Gene-sets

We investigated the burden of PTCs within CHD patients by examining their distribution among gene-sets annotated by Gene Ontology Biological Processes (GO:BP) and Molecular Function (GO:MF), KEGG, and Reactome using the SKAT package (see methods).^18^ We adjusted for ancestry, and set FDR at 0.05 for each analysis. For significant GO:BP gene-sets, we focused on the subset of “developmental processes” family of terms (GO:0032502; Additional File 2: Data 1 and 2). Significant terms recovered were then grouped based as to whether the process contributes to development broadly, or specifically to the heart, the brain, or any other individual organ. All PTCs combined (PTCall) from CHD patients fall disproportionately into heart and brain development related terms (Table 1). The high proportion of brain development terms supports mounting evidence for the sharing of genes regulating heart and brain development.^19–22^ When this analysis was repeated separately for PTCs stratified by NMD annotation (NMDtrig or NMDesc), as expected, a large fraction of both the NMDtrig and NMDesc PTCs were associated with heart development gene-sets. Interestingly, while the fraction of significant heart development terms were not significantly different for PTCall and NMDtrig PTCs (26% vs 22%), a significantly higher proportion was observed for NMDesc PTCs (47%; p = 0.04 compared to PTCall and p = 0.02 compared to NMDtrig). This suggests NMDesc PTCs may play a disproportionate role in heart specific developmental pathogenesis of the heart in CHD—as opposed to disrupting both cardiac and extracardial development.

**Table 1.** Predicted NMD for GO:BP Developmental Process Classified by Organ.

|  | Any | Heart | Brain | Other |
| --- | --- | --- | --- | --- |
| All PTCs (n=171) | 49<br>(29%) | 44<br>(26%) | 31<br>(18%) | 47<br>(27%) |
| NMDtrig (n=134) | 45<br>(34%) | 30<br>(22%) | 26<br>(19%) | 33<br>(25%) |
| NMDesc (n=15) |  |  |  |  |
| Observed | 6<br>(40%) | 7*<br>(47%) | 1<br>(7%) | 1<br>(7%) |
| Expected | 4.30 | 3.86 | 2.72 | 4.05 |

Distribution of PTCs within GO “developmental processes” gene-sets categorized as either being non-specific (any), specific to heart development, brain development, or the development of any other organ (other). \*Statistically significant based on 2 population z-score tests compared to either All PTCs ( $p = 0.04$ ) or to NMDtrig ( $p=0.05$ ).

### Processes and Pathways Identified by NMDesc and NMDtrig PTCs

Investigating the heart development terms that were recovered from the GO:BP analysis showed a burden of PTCs within pathways related to heart valve development. While both NMDtrig and NMDesc PTCs recovered the parent term “Heart Valve Development” (Additional File 1: Table S2), the NMDtrig variants were also significant only for “Aortic Valve Morphogenesis”, while NMDesc variants were also significant only for “Atrioventricular Valve Development”, suggesting differing genes with differing mechanisms of disruption. The NMDesc PTCs in the significant terms fell within genes such as *BMP2/BMPR1a,* consistent with the known requirement for BMP signaling in atrioventricular valve development,^23–2526^ while NMDtrig PTCs were found in *ELN, NOS3, JAG1/DLL3,* genes known to regulate aortic valve development. It is notable that *ELN* encoding the matrix protein elastin is linked with William syndrome, a genetic disorder characterized by supravalvular aortic stenosis. Also observed among NMDtrig PTCs is another atrioventricular valve disease gene, *DSCH1,* associated with mitral valve prolapse (Additional File 2: Data 3).^27^ Also recovered with NMDtrig PTCs were several genes (*SNAI1, NFATC1, TGFB)* regulating epithelial-mesenchymal transition (EMT), which are critical for valve morphogenesis.

Turning our eyes to the analysis of pathways annotated by GO Molecular Function (GO:MF), KEGG and Reactome gene-sets, NMDesc PTCs recovered 15 pathways (12 shown in Figure 2). Of these, 7 were also significant when testing NMDtrig PTCs (Additional File 2: Data 4, 5, 6). It is notable that “MAPK signaling” was exclusively associated with NMDesc PTCs (Figure 2b; KEGG). Dominant or de novo gain-of-function (GOF) variants in the RAS/MAPK pathway are well described to cause Noonan syndrome, a syndromic disorder often accompanied by right ventricular outflow obstruction associated with pulmonary stenosis.^16,17^ Consistent with this, two genes (*SOS2, KRAS*) known to cause Noonan syndrome were recovered housing NMDesc PTCs, although none of the CHD patients in this study are known to have Noonan syndrome (Additional File 2: Data 7).

**Figure 2.**
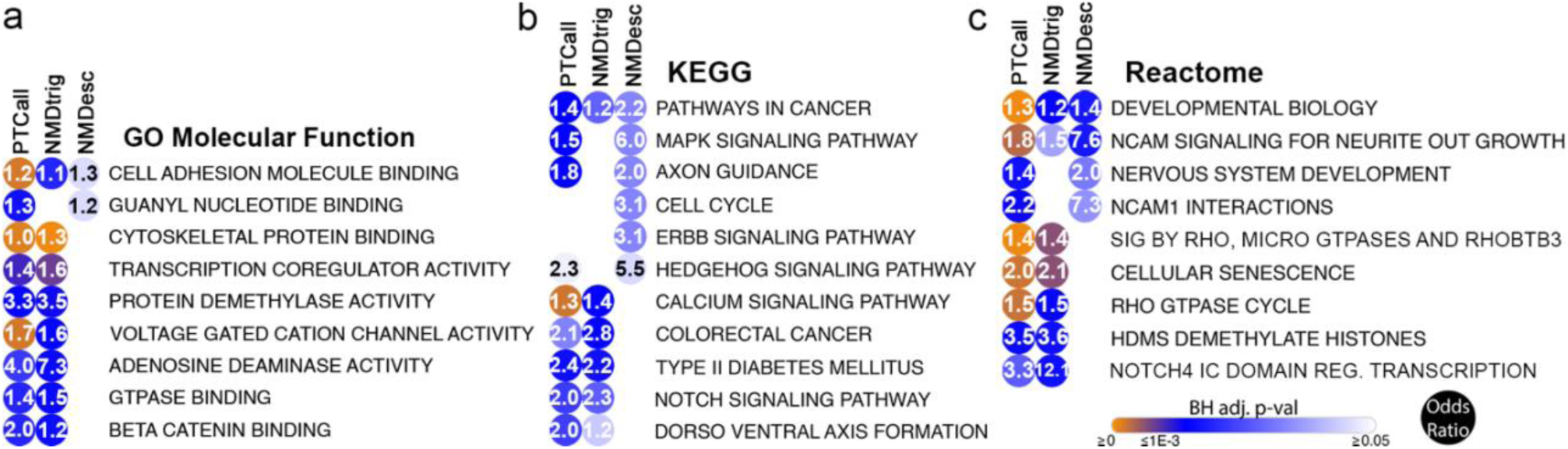
Distribution of PTCs within Processes and Pathways Differ by predicted NMD interaction. All PTCs (PTCall), the subset of NMDesc PTCs, or the subset of NMDtrig PTCs show are significant within a different distribution of gene-sets complied by (**a**) Gene Ontology (GO) Molecular Function, (**b**) Kyoto Encyclopedia of Genes and Genomes (KEGG) or (**c**) Reactome biological databases.

Among pathways exclusively associated with NMDesc PTCs are Hedgehog and ERBB signaling (Additional File 2: Data 8). Genes recovered with PTCs in the Hedgehog pathway included *GLI1, GLI2,* and *SUFU.* Both of these cell signaling pathways are well described to cause CHD in mouse models, but there is a paucity of evidence for their contribution in human CHD. Other pathways exclusively recovered as associated with NMDtrig PTCs were “Notch signaling” in KEGG and the related “Notch4 IC domain regulated transcription” in Reactome (Figure 2c). Multiple studies have previously shown the association of *NOTCH1* variants with aortic valve defects and other left ventricular outflow tract CHD.^28,29^

The burden analysis of PTCs using gene-sets also recovered multiple pathways related to guanyl nucleotide signaling, and particularly GTPase activity (Additional File 1: Table S3). For example in GO:MF, “Guanine Nucleotide Binding” was associated with NMDesc, while “GTPase binding”was associated with NMDtrig. In Reactome, “Signaling by Rho, microGTPases, and RhoBTB3” and “Rho GTPase Cycle” were both associated with NMDtrig. These findings suggest a possible distribution of positive regulation in GTPase signaling.

### NMD Transcript- and Gene- Level Burden Analysis Recovered Novel CHD Genes

Genome-wide transcript- and gene- level burden analysis using predicted NMDtrig and NMDesc PTCs yielded 11 candidate CHD genes, 3 from analysis of NMDtrig variants (*ABDH11, KDM6B*, and *FLT4*), and 8 from analysis of NMDesc variants (*ZNF792, ZNF77*, *BSN*, *RERE*, *ZDHHC8*, *CACNA1G, TRIM59*, *MATN4)* (Table 2, Additional File 4: Supplemental Figure 1). Among the 11 genes, *RERE* and *FLT4* were previously curated as CHD causing among a list of 143 high confidence CHD genes, yielding significant enrichment for known CHD genes among our 11 recovered genes from analysis of NMDtrig and NMDesc variants (hypergeometric test, p = 0.003).^30^

**Table 2.**
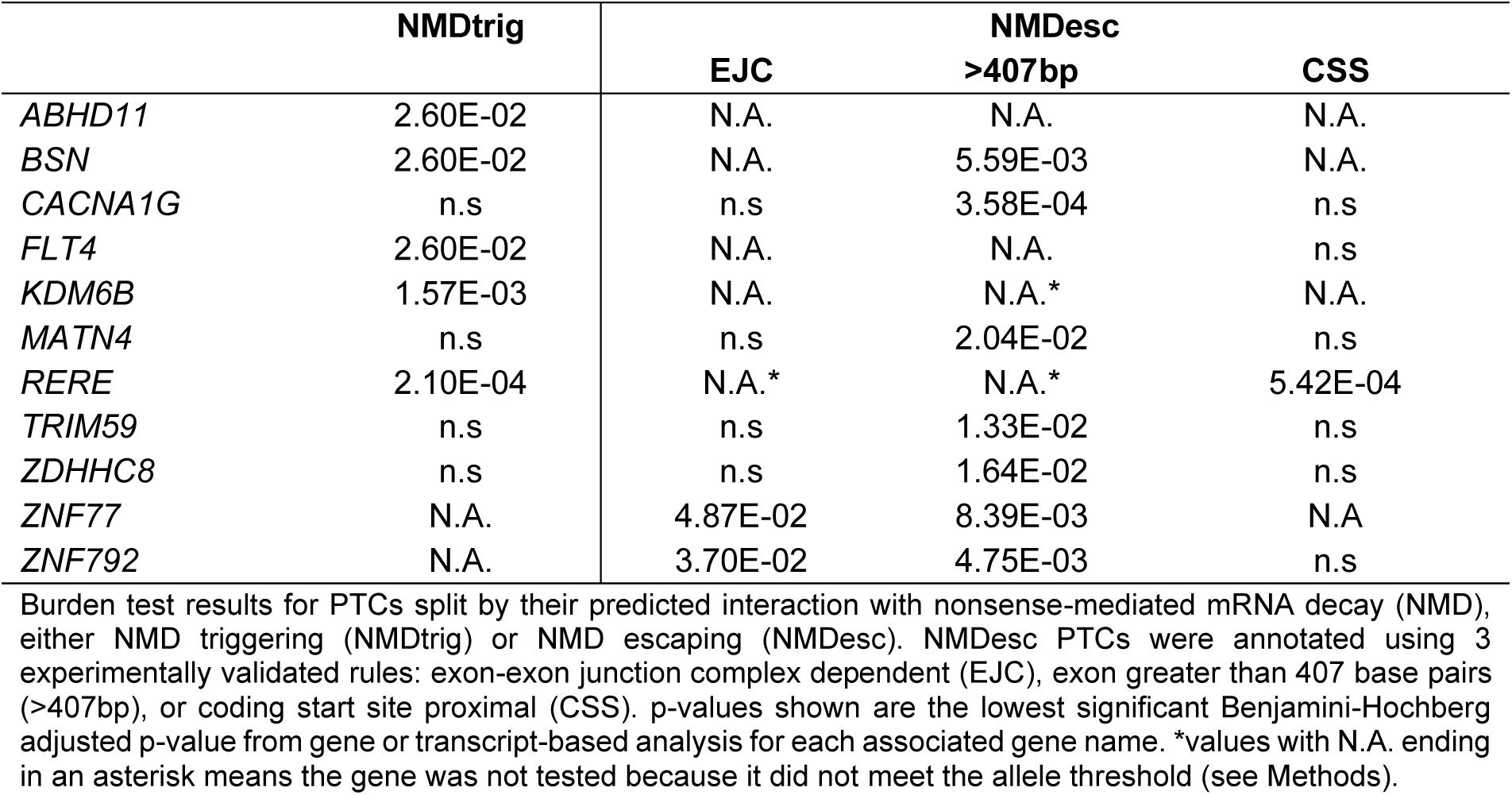
Eleven Loci Associated with CHD Identified Using Burden Analysis of NMDtrig or NMDesc PTCs.

Of the 3 genes recovered from analysis of NMDtrig variants, neither *ABDH11,* abhydrolase domain 11, nor *KDM6B,* lysine demethylase 6B, were associated with CHD, although *ABDH11* is on human chromosome 7q11.23, a region when deleted can give rise to William Beuren syndrome associated with aortic defects.^31^ For *KDM6B,* analysis of de novo PTVs from clinical patients showed a 14% incidence of CHD.^32^ The third gene recovered, *FLT4*, is a receptor tyrosine kinase previously associated with Tetralogy of Fallot (TOF), a CHD constellation of four individual cardiac lesions. In addition to the previously reported PTCs from the PCGC cohort, another PCGC patient and a CHP patient were found with previously reported PTCs. Both Heterozygous inherited and *de novo* variants, assumed to cause LOF-induced haploinsufficiency from NMDtrig PTCs, were associated with TOF from at least two studies.^20,33^

For NMDesc variants, 8 genes were recovered as significantly associated with CHD, including: *ZNF792, ZNF77*, *BSN*, *RERE*, *ZDHHC8*, *CACNA1G, TRIM59*, *MATN4* (Figure 3). All were recovered as heterozygous variants, except for one homozygous variant in *TRIM59*. LVOTO was the most common phenotype, observed with 5 of the 8 genes (Additional File 1: Table S4). Interestingly, *BSN* (Bassoon presynaptic cytomatrix protein; Figure 3b) and *RERE* (atrophin family member with transcriptional repressor activity; Figure 3c) are predominantly expressed in the brain (Additional File 1: Table S5). Only *RERE* is known to be associated with CHD via the disorder NEDBEH, Neurodevelopmental Disorder with Anomalies of the Brain, Eye, or Heart.^34,35^ Of the 14 patients with *RERE* variants, 4 had atrial septal (ASD) and ventricular septal defects (VSD), and 9 had valvular anomalies, phenotypes aligned with that of NEDBEH and the observed role of *RERE* in atrioventricular valve development (Additional File 3: RERE).^36^ For ZDHHC8, a transmembrane protein with predicted palmitoyltransferase activity, all the NMDesc PTCs were located beyond the catalytic domain, near the intracellular N-terminus with the regulatory and binding regions of this class of proteins, (Figure 3d).^37^ Interestingly, 5 patients have 2 different frameshift variants causing a PTC at the same position (Figure 3e), suggesting this may have functional relevance, perhaps via dominant effects mediated by a N-terminal truncated ZDHHC8 protein.

**Figure 3.**
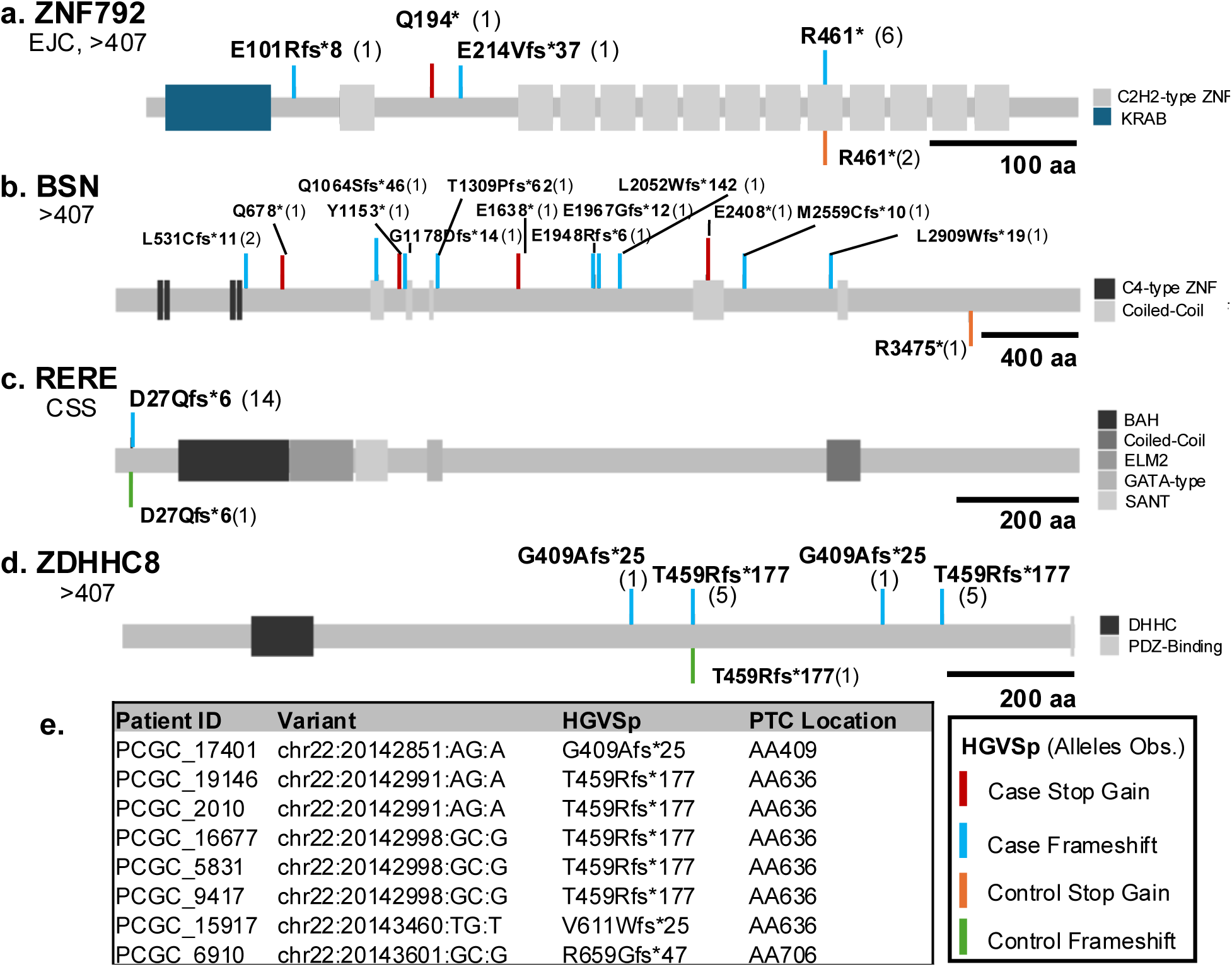
PTCs in recovered CHD-associated Genes and Their Protein Impact. In diagrams (**a - d**), the gene symbol is listed with the corresponding rule that caused the loci to reach significance. Each lollipop represents a unique PTC-inducing variant and the corresponding HGVS protein annotation, with the number of alleles in cases or controls being enclosed within parentheses. (**e**) A table of CHD cases with a NMDesc PTC in ZDHHC8, their corresponding genomic variant and HGVS annotation, as well as the amino acid (AA) location of the downstream truncation. Protein structure information was obtained from Uniprot and Interpro.

CACNA1G, encoding a CaV3.1 T-type voltage-dependent calcium channel, was a notable result, as it was found with PTCs localized to two different exons longer than 407bp; one exon near the 5’ end and the other at the 3’ end of the transcript (Additional File 4: Figure 1). If translated, NMDesc of transcript with 5’ PTCs may lead to loss of function, while the 3’ PTCs may exert dominant effects. This spread of PTCs to the 5’ and 3’ end may account for this gene being overlooked, as the PTC at the final exon would have been filtered out.

### Role in Developmental Disorders and Regulation of Cell Proliferation and Cell Signaling

We further investigated the 11 CHD genes recovered from the NMD analysis using Ingenuity Pathway Analysis (IPA) for insights into possible underlying disease mechanisms. IPA analysis using direct interactions yielded a 35-gene interactome network that incorporated all 11 of our recovered CHD genes (Figure 4a). IPA enrichment analysis yielded an annotation of “developmental disorder, hereditary disorder, and neurological diseases.” Interestingly, a large part of the nodes linked to the 11 input genes in this network are ubiquitination-related (UBC, CUL3; black asterisks), while a smaller portion is involved in membrane dynamics (ARF6, PLEKHA4, TRIM67; black caret). The latter is reminiscent of the recovery of multiple pathways related to membrane and cytoskeletal dynamics in the gene-set analyses above (Figure 2). Further interrogation of the top enriched terms in this analysis identified Moebius syndrome—a birth defect with high rates of CHD (33%) co-occurring with intellectual disability (p = 1.03E-5; *BSN*, *KDM6B*)(Additional File 1: Table S6).^38^ Also recovered were congenital neurological disorder (p = 2.25E-4; *BSN*, *KDM6B*, *CACNA1G*, *MATN4*, *RERE*) and growth of vessels (p=4.74E-4; *CACNA1G*, *FLT4*, *KDM6B*).

**Figure 4.**
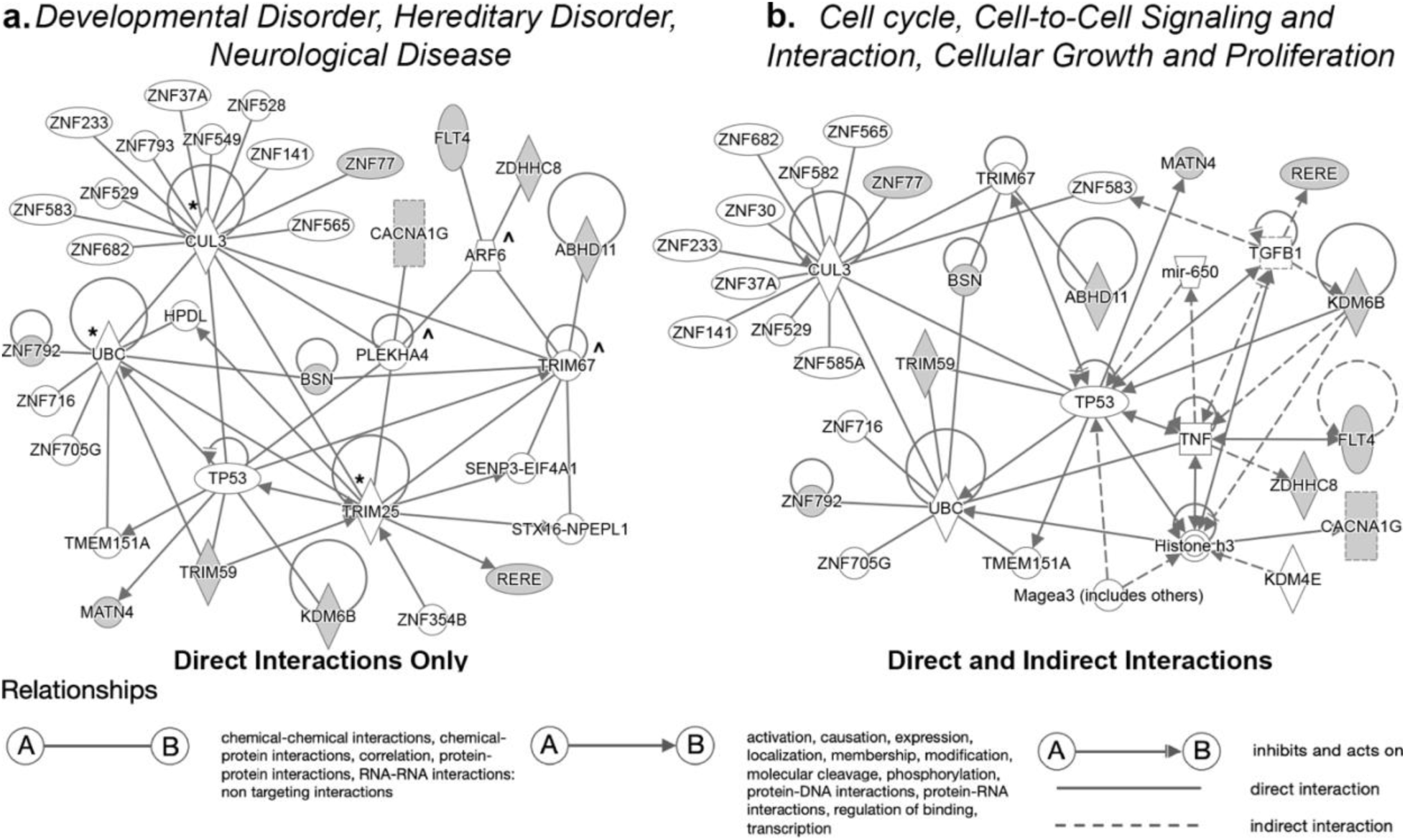
IPA Interaction Networks from the 11 CHD Genes Recovered from NMD PTC analysis. Biologically annotated networks created with the 11 significant genes (grey nodes) using either (**a**) direct, or (**b**) direct and indirect interactors. More detailed key available online via <u>Qiagen</u>.

Expanding the IPA analysis to include both direct and indirect interactions yielded another compelling network that similarly incorporated all 11 CHD genes in a 35-gene network. This network was annotated as “cell cycle, cell-to-cell signaling and interaction, cellular growth and proliferation” (Figure 4b). Prominent linking nodes comprise Tumor Necrosis Factor (TNF), Transforming Growth Factor β1 (TGFB1) and TP53 (tumor protein P53), genes with essential functions in cell proliferation and cell signaling, and in many processes regulating cardiovascular development and function. These findings are reminiscent of the above pathway enrichment analysis that recovered related pathways such as “cell cycle”, “pathways in cancer”, “cytoskeletal rearrangement”, and “senescence” (Figure 2). Together these findings support the 11 CHD genes as being functionally related and part of a larger interactome network contributing to CHD pathogenesis via the regulation of biological and cellular processes important in heart (and brain) development.

### Protein-Protein Interactome Analysis Show CHD Gene Enrichment

To explore whether direct protein-protein interactions (PPI) may serve as a context for CHD pathogenesis, our 11 recovered CHD genes were examined for first order PPIs. This recovered 226 primary interactors, among which are 8 additional suspected CHD genes that are also found among a list of 143 high confidence CHD genes.^30^ This corresponds to a significant enrichment for CHD genes among the primary interactors of the PPI network (hypergeometric test; p = 0.0002) and supports the notion that the network has a functional relevance in the pathogenesis of CHD (Figure 5; Additional File 4: Figure 2). To further assess the functional importance of PPI networks in CHD, we used the sequence kernel association test (see methods) to assess the association of rare pathogenic variants within the 11 CHD genes and their 226 primary interactors with CHD. For this analysis, a broader range of predicted pathogenic variants were included in this analysis including missense, in-frame indel, and protein truncating variants. This analysis yielded 5 significant genes, including 3 genes recovered in the PTC analysis—*ABHD11 (*BHadj-pval = 0007*), KDM6B* (BHadj-pval = 0.001) and *TRIM59* (BHadj-pval = 0.007). Also recovered are two other genes, *ERBB2* (BHadj-pval = 0.007) and *TRIM41* (BHadj-pval = 3.24E- 5) (Additional File 2: Data 9). The recovery of *ERBB2* is notable, as ERBB signaling was recovered in the analysis of the NMDesc PTCs (Figure 2b). We note a recent study reported *ERBB2* missense variant associated with LVOTO lesions.^39,40^ The recovery of *TRIM41* (3 recurring in-frame deletions) and *TRIM59* (NMDesc variants) are interesting, as their role in CHD pathogenesis has not been reported even as there is increasing interest in the TRIM family of proteins in other cardiovascular diseases.^41^ Together these findings suggest the CHD-interactome may provide a functional framework for interrogating the genetic etiology of CHD.

**Figure 5.**
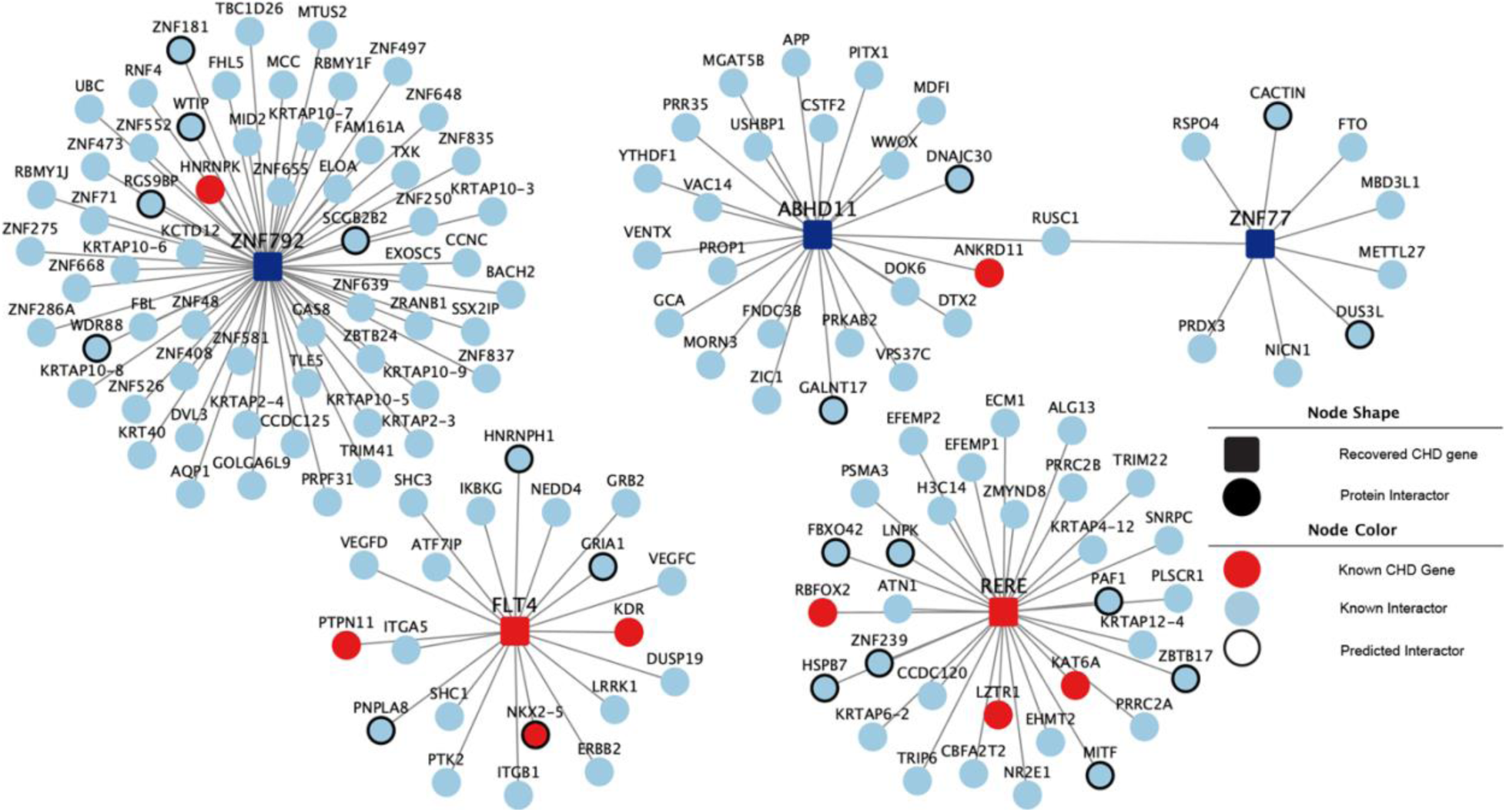
Primary Protein-Protein Interactions Generated by 5 of the 11 CHD genes recovered by NMD PTC analysis. Shown as the central nodes are 5 of our 11 CHD genes recovered from NMD PTC analysis and their primary protein-protein interactors. High-confidence CHD genes found in the database CHDgene are further highlighted in red. Interactions for all 11 genes can be found in Additional File 4: Figure 2.

### Digenic and Oligogenic Interactions in CHD Pathogenesis

Analysis of the NMDtrig and NMDesc PTCs that yielded the 11 CHD-associated genes showed an unexpected cooccurrence of these PTCs in digenic pairs within patients. PTCs in 9 unique digenic pairs between 9 of the 11 CHD genes were observed in 9 CHD patients (Figure 6). Based on the per gene frequencies of PTCs and the size of our patient cohort, random chance would have predicted only 4.2 patients with such digenic pairs, suggesting that these digenic pairs may have disease relevance. To further assess the potential contribution of digenic combinations of PTC variants in CHD pathogenesis, we queried for the co-occurrence of PTCs for genes in the heart development pathways recovered in our GO:BP analysis above (Additional File 2: Data 2). This yielded nearly twice the proportion of CHD patients with more than one PTC variant vs. control subjects (90/600= 15% vs 58/766=7.6%; two-sided proportion test, p-val = 3.851e-05). In contrast, querying gene-sets from the totality of the 171 GO:BP developmental process terms yielded no such significance, with 66.7% observed in CHD patients and 64.5% in control subjects. As our CHD subjects have a larger number of PTC alleles per person as compared to the control subjects (9.46 vs 8.88 alleles, p = 1.347e-08), the enrichment is likely to be underestimated. Overall, these findings are in line with the higher frequency observed for digenic pairs with PTCs for the 11 CHD genes in CHD patients. We then analyzed the digenic, trigenic, and quadgenic combinations of genes with PTCs in the GO:BP heart development curated gene-sets. *KDM6B* stood out as the most frequently reoccurring gene (14), followed by *CACNA1G* (6), *MEGF8* (5) *and NOTCH3* (4) (Figure 7; Additional File 4: Figure 3). It is notable that there were no recurring gene pairs other than NOTCH3-XIRP2, which was observed in two patients. These findings suggest many genes in different gene combinations can contribute to the pathogenesis of CHD.

**Figure 6.**
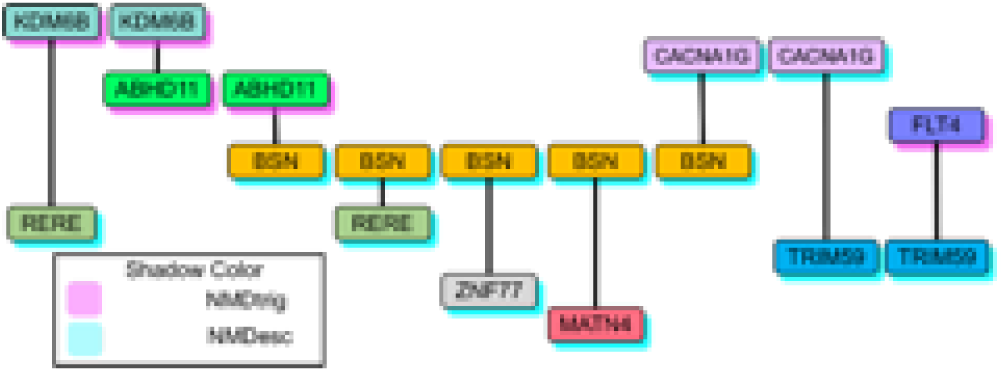
Digenic Combinations of PTCs among 9 Recovered CHD genes. Nine non-repeating digenic pairs of PTCs recovered from patients involving 9 of 11 total recovered CHD genes.

**Figure 7.**
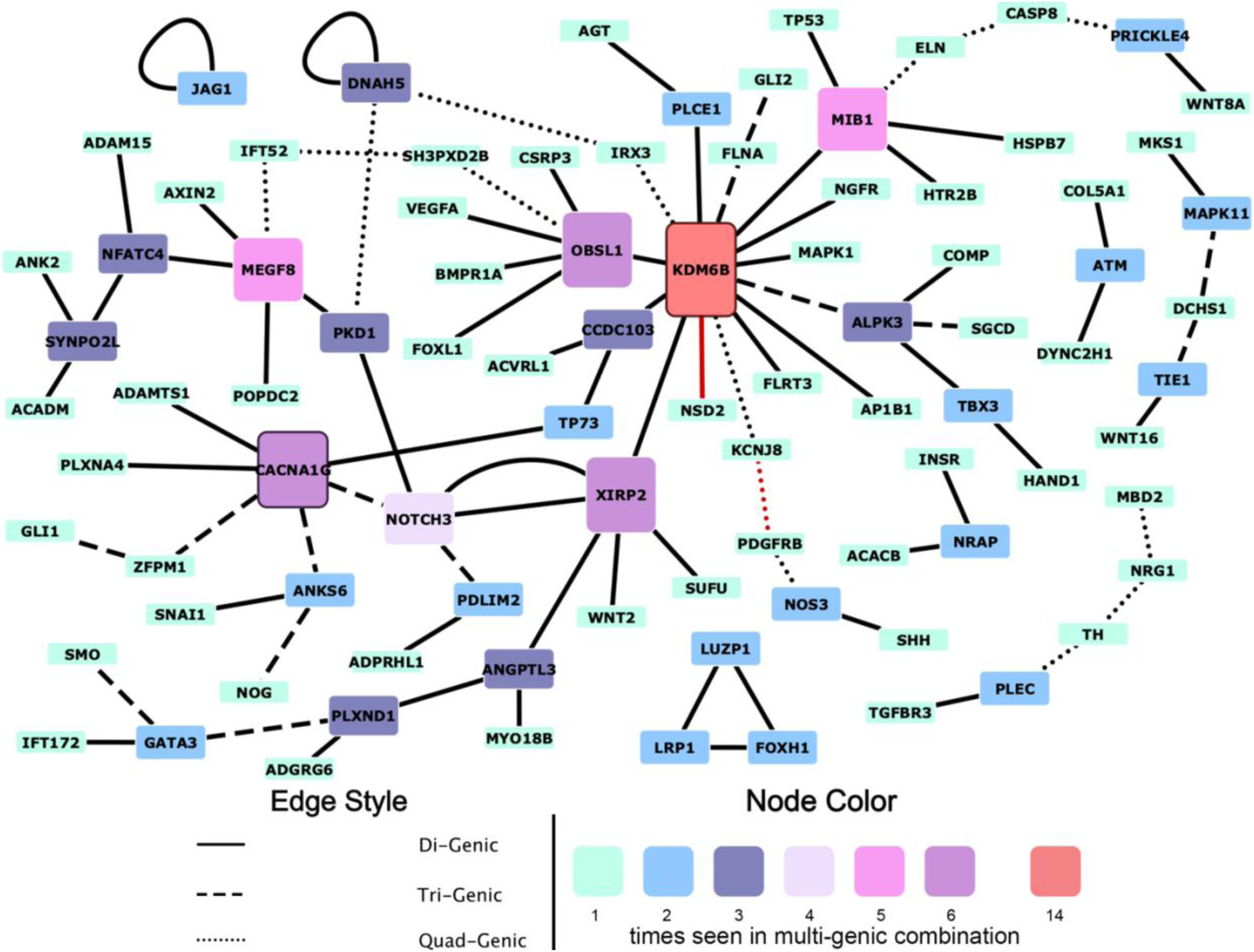
Oligogenic Combinations of PTCs in Heart Development Genes found in CHD patients. Heart development associated gene-sets previously recovered from analysis of PTCs were further interrogated and found to have increased incidence of oligogenic occurrence, observing digenic, trigenic and quad-genic combinations. Each combination was unique except for two patients with a NOTCH3-XIRP2 PTC pair. Notable is KDM6B, seen in 14 unique oligogenic sets corresponding to 14 patients. A subset of PTC pairs from non-repeating genes was excluded here and can be found in Additional File 4: Figure 3.

## DISCUSSION

The analysis of human exome sequencing data has typically been based on the association of *de novo* variants predicted to be LOF and have thus far yielded significant insights into the genetic etiology of CHD. These studies analyzing predicted LOF variants have included NMDtrig PTCs, but NMDesc PTCs have not been examined systematically and are even discarded in other studies. In the present study, we conducted a systematic analysis of all PTCs, stratified by their predicted interactions with NMD, including those that are predicted to escape from NMD. Transcripts with NMDesc PTCs have the potential to encode anomalous protein products that can exert dominant effects. Our analysis of PTCs showed a significantly larger fraction of NMDesc PTCs fall within gene-sets directly related to heart development as compared to NMDtrig PTCs, indicating that NMDesc PTCs may play a disproportionately greater role in the developmental pathogenesis of CHD.

Among heart development terms, significant association was observed for heart valve development for both NMDtrig and NMDesc PTCs. While NMDtrig PTCs were only associated with the aortic valve development gene-set, atrioventricular development was observed as significant for NMDesc PTCs. Analyzing our NMDtrig PTC pathway gene-set results terms related to Notch signaling were recovered, a cell signaling pathway essential for aortic valve development. NMDtrig variants were recovered in *ELN* and *JAG1,* genes associated with aortic valve defects in William syndrome and Alagille syndrome, respectively, as well as *NOS3*, a gene associated with bicuspid aortic valve. In contrast, analysis of the NMDesc PTCs recovered BMP signaling, a pathway critical in atrioventricular valve development. Aligning with this, NMDesc PTCs were recovered in a gene linked to mitral valve prolapse, such as *DSCH1*. NMDesc PTCs also were recovered in genes regulating EMT, a process regulating formation of the endocardial cushion that gives rise to both atrioventricular valves.

Particularly notable was the recovery of Hedgehog and ERBB signaling exclusively with NMDesc PTCs. These genes can cause a wide spectrum of CHD in mice, yet human studies to date have not confirmed their role in CHD pathogenesis except in syndromic disorders, such as in some instances of Carpenter’s syndrome—associated with dysregulated sonic hedgehog (SHH) signaling—or syndromic neurological/neurodevelopmental disorder—associated with ERRB signaling.^42–46^ Supporting a role for ERBB signaling in CHD pathogenesis is a report detailing an *ERBB2* variant associated with left ventricular outflow obstructions.^47^

Also observed as exclusively associated with NMDesc PTCs is “MAPK signaling”, a pathway in which dominant GOF variants are known to cause Noonan syndrome.^16,17^ As none of our CHD patients have Noonan syndrome, this suggests the possibility that disturbance of this pathway may also contribute to nonsyndromic CHD. Interestingly, we observed multiple guanyl nucleotide related gene-sets, including Rho and Ras GTPase, which were recovered by either NMDesc or NMDtrig PTCs. These enzymes are known to have critical roles in regulating development of the cardiac outflow tract and endocardial EMT through their role in cytoskeletal and membrane dynamics.^48,49^ NMDesc PTCs were observed as associated with guanine nucleotide binding and NMDtrig with GTPase activity, findings that could suggest their roles in disrupting positive regulation of cardiac development. This is consistent with the known positive regulation of GTPase signaling by GEFs (guanine exchange factors).

Our gene-level analysis yielded 11 genes associated with CHD, 9 from analysis of NMDesc PTCs, and all except for *RERE* and *FLT4* being novel genes not previously known to cause CHD. The recovery of *CACNA1G*, a gene encoding a CaV3.1 T-type voltage-dependent calcium channel, is particularly notable. Two groups of NMDesc PTCs were recovered in this gene, one toward the 5’ end between regions encoding ion transport domains and the other group localized toward the 3’ end in the intracellular regulatory domain region. While the 5’ NMDesc domains would lead to loss of function, the 3’ PTCs could exert dominant effects if expressed into an anomalous protein. This exemplifies how PTCs, depending on their location and the gene exon structure, may exhibit different molecular mechanisms, routes of inheritance, and clinical manifestations.^12,50,51^ We note significant risk for lethal arrhythmias are observed with CHDs, particularly those with single ventricle defects.^52^ Interestingly, we observed 4 of the 11 CHD patients with CACNA1G NMDesc PTCs have single ventricle CHD, 3 being HLHS (p = 0.068). In patients with HLHS, an enrichment of de novo variants in conduction system genes have been recently reported.^22^ Together these findings suggest the high risk of arrhythmias in single ventricle patients may have a genetic underpinning.

We observed the 11 CHD genes and their primary protein interactors show enrichment for high-confidence CHD genes, supporting relevance of the 11 genes recovered from the NMD analysis in CHD pathogenesis. IPA of the 11 CHD genes yielded two compelling interactome networks, one annotated as “developmental disorder, hereditary disorder, and neurological diseases” and the other network annotated as “cell cycle, cell-to-cell signaling and interaction, cellular growth and proliferation.” Together, these findings suggest interactome networks may provide the genomic framework defining the genetic complexity of CHD. This idea has begun receiving attention not only in CHD but also other diseases. In Bardet-Biedl syndrome, which also can be accompanied by CHD, digenic pairs of damaging variants in distinct networks of ciliary proteins are thought to drive disease.^53^ An interactome network of cilia genes also have been observed in the context of CHD,^54^ but other protein complexes (e.g. chromatin modifiers) and biological networks (e.g. “brain connectome”) have also received attention.^55^

Our findings support a digenic/oligogenic model of disease, as unique digenic pairs of PTCs within the 11 CHD genes were recovered in 9 CHD patients. Also noted was the proportion of CHD patients with more than one PTC variant in genes from significant heart development pathway gene-sets, an observation not seen when considering all development-associated genesets. This included higher order combinations of genes with PTCs beyond the digenic pairs, including trigenic, and quadgenic combinations. Alongside the observation that *RERE* mouse model phenotypes depend on genetic background and accompanying variants, and similar reports from other CHD mouse models, these findings suggest a role for oligogenic etiologies in the pathogenesis of CHD.^36,56–61^ Based on these findings, we propose that genetic interaction networks may provide the genomic framework in which the pathogenesis of CHD emerges. This could account for the high prevalence of CHD, which is seen at an estimated incidence of 0.5%, and contribute to the complex genetic and seemingly sporadic nature of CHD.

Overall, findings from our PTC variant analysis yielded evidence supporting the importance of NMDesc PTCs in the pathogenesis of CHD. These may have disproportionately greater roles in CHD pathogenesis. Analysis of both NMDtrig and NMDesc variants have uncovered new genes and pathways likely contributing to human CHD. Our findings provide support for NMDesc PTCs with possible GOF effects in the MAPK and Rho GTPase signaling contributing to the genetic etiology of nonsyndromic CHD. The verification of these findings in future studies will require experimentally confirming the downstream impact of NMDtrig/NMDesc PTCs using transcript and protein expression analysis from patient cells/tissues paired to genomic sequencing data.

## Methods

### 1.1.1 Whole exome sequencing analysis

For 656 subjects recruited at the Children’s Hospital of Pittsburgh UPMC (CHP), whole exome sequencing (WES) sequencing was carried out on Illumina HiSeq2000 with 100 paired-end reads at 80-100X coverage using Agilent V4 or V5 exome capture kit. For 2425 samples obtained from the Pediatric Cardiac Genomics Consortium54 (PCGC, dbGaP phs001194.v2.p2) SRA files were downloaded from the NCBI SRA database, and for 5140 healthy control samples obtained from the Alzheimer’s Disease Sequencing Project (ADSP, NG00067.v2) (with no personal or family history of dementia-related disease), gVCFs were obtained from the National Institute on Aging Genetics of Alzheimer’s Disease Data Storage Site. Alignment of reads was done using the human reference genome GRCh38. The entire germline variant calling, joint-calling, variant QC, and sample QC was conducted as previously outlined (Williams, 2022), with the exception that no samples were removed based on ancestry. Principal Component Analysis (PCA) was conducted for the entire cohort, and the first 4 principal components were used to account for differences in ancestry.

Even though the overall distribution of LOF variants (stop gain, frameshift, splice site) was the same between the three cohorts, controls displayed a higher average of PTC alleles per sample than cases (9.45 control vs 8.88 cases, t-test p-value = 1.37e-8; Additional File 1: Table S1). These variants also appeared less frequently, with both the average in-cohort frequency of these variants (3.56 vs 3.86, Wilcoxon rank p-value < 0.0066) and gnomAD average allele count (29.38 vs 35.64, Wilcoxon rank sum p-value = 2.44e-8) being lower in controls than in cases. Initially, we presumed this may be due to the difference in age of the cohorts – the patients in the CHD cohorts were sequenced at an early age while control patients from ADSP were sequenced at an old age –, as the phenomenon of clonal expansion of indeterminate potential (CHIP) is linked to a raise in PTC within a specific group of genes. However, after removing all PTCs in the top 11 CHIP genes from both cohorts the difference persisted.

### 1.1.2 AENMD Annotation of Genomic Variants and Transcript-to-Gene-to-Gene-set Consolidation

The joint called and QC filtered VCF was read in and processed by AENMD v0.3.11 paired with aenmd data package v0.3.2, which utilizes ensembl release version 105 for annotation (Klonowski, Liang, Coban-Akdemir, Lo, & Kostka, 2023). Default settings were used for the prediction of PTC-transcript pairs’ sensitivity to NMD detection (d_pen = 50 and d_css = 150) and only using high confidence (TSL = 1) or single exon/intronless transcripts (TSL = NA). The results from AENMD’s last coding exon and d_pen rules were grouped under the canonical exon– exon junction complex dependent model (EJC). Further, all PTC-transcript pairs in single coding exon and intronless transcript were removed when testing the EJC rule because AENMD incorrectly labeled these as NMDesc based on the last coding exon rule. A results table summarizing the PTC-transcript annotation statistics for each of the 3 rules can be found in Additional File 1: Table S7. Next, in preparation for gene-set and gene-level analysis, PTC-transcript pairs were consolidated to provide a single annotation per PTC-associated gene model. Briefly, for PTC-transcript pairs mapping to the same gene the selected NMD annotation was selected based on the criteria (1) MANE Select transcript’s call, (2) the Ensembl canonical transcript’s call, (3) majority rule, and (4) if no majority is found, NMDesc is selected. A results table containing summarizing PTC-gene pair annotation statistics for each of the 3 rules can be found in Additional File 1: Table S8.

### 1.1.3 Gene-set Annotation of Genomic Variants

Gene-sets compiled by gsea-msigdb (gsea-msigdb.org) were downloaded using the msigdbr v7.5.1 R package (accessed on 28/08/2023) (Dolgalev, 2022). For each gene-set source (species = human, category = C2 or C5, subcategory = KEGG, REACTOME, GO:BP, GO:MF), PTCs were assigned to all applicable gene-sets under each source (e.g. GO:BP’s actin_cytoskeleton_reorganization) based on the ENSG that they were mapped to by AENMD. Of note is that PTCs were allowed to be mapped to multiple ENSGs and the same PTC-ENSG pair could be mapped to multiple gene-sets under the same source (e.g. GO:BP’s actin_cytoskeleton_reorganization and actin_filament_based_movement).

### 1.1.4 SKAT Burden Analysis

PTCs grouped by gene-set, gene, or transcript were analyzed under the burden framework within SKAT v2.2.5 (Wu et al., 2011). The null model, SKAT_Null_Model(), was established using the first 4 principal components as covariates to account for ancestry, as well as a covariate for reported sex. Further, the out_type was set to “D” for dichotomous outcome (CHD or no CHD), n.Resampling set to 2000, and type.Resampling as “bootstrap.” The robust SKAT procedure for binary outcomes, SKATBinary_Robust.SSD.All(), command with the method set to “Burden” was used for all analyses (Zhao et al., 2020). A summary of the SKAT analysis may be found in Supplemental Workbook 2 for the gene-set (Additional File 1: Table S9), gene (Additional File 1: Table S10) and transcript (Additional File 1: Table S11) analysis. Results from gene-set analysis may be found in Additional Files 2 Data 1, 4-6, with p-values being adjusted for false discovery rate using the Benjamini Hochberg correction (FDR-BH) and odds ratio calculated by a classic 2×2 table analysis – note that the odds ratios are estimates as the covariate structure cannot be accounted for in their calculation. Before viewing the results, gene and transcript findings were subjected to an adaptive minor allele count (MAC) threshold on a per rule basis, such that all genes and transcripts without sufficient alleles were removed. The thresholds used were as follows: 9 MAC for EJC, 9 MAC for >407, and 13 for CSS. After removing results under the threshold, FDR-BH was used to adjust p-values. For transcript and gene level results, a complete set of significant results including raw p-values, adjusted p-values and odds ratios may be found in Additional File 2: Data 10-12. Original SKAT output can be found in <u>Additional</u> File 5. In the gene and transcript results, there were four significantly associated genes that we did not report on in the body of the text (AOC2, DNMT3A, TET2, ASXL1). Upon expectation of these three results, all were enriched in controls. While stands to reason that AOC2 EJC NMDesc are possible candidate protective alleles, we realized that DNMT3A, TET2 and ASXL1 are artifacts arising from the clonal hematopoiesis of indeterminate potential (CHIP) phenomenon (Marnell, Bick, & Natarajan, 2021). In addition to checking for evidence for brain expression for our significant genes from this analysis, we assessed these genes for adult heart expression, developmental heart expression, and their predicted sensitivity to allele loss (Additional File 1: Table S12-14). Finally, all the patients with PTCs in the significant genes, their phenotype classification and phenotypes can be found in Additional File 3.

In the instance of testing for association between CHD and the interactome of 237 genes (our 11 CHD genes and their 226 primary interactors), we used a modified version of the procedure used above. The first difference is that we used rare (<1%), deleterious missense (as predicted by REVEL or ESM1B), inframe indel, and high impact (as defined as Ensembl) variants, grouped by gene, instead of PTC-inducing variants. The remaining steps were the same, except that the method set for the SKATBinary_Robust.SSD.All() command was “SKAT” instead of “Burden.” We used this test because, despite our best efforts, we expect that variants within the same gene may have different, and even opposing, effects on phenotype. The presence of variants with differing effects (i.e. positive, neutral, or negative effects) when using the burden method drastically lowers the power of this test. SKAT is a variance component test that accounts for these differences by testing for overdispersion, or an increase in variance, of the distribution of variants in a gene between cases and controls.

### 1.1.5 Gene Ontology Biological Processes Ontology Analysis

Analysis of the Biological Processes Gene Ontology (GO:BP) Ontology was conducted using the ontologyIndex v2.11 R package using the go_2023-01-01.obo file from the Gene Ontology Data Archive at http://release.geneontology.org/ (Greene, Richardson, & Turro, 2017). All significant results from the GO:BP that were child terms to GO ID 0032502 were then analyzed further. For each of the 171 terms, the term was classified as either participating in a general developmental process (any), specific to heart development (heart), specific to the cardiovascular system, specific to the brain (brain), specific to the peripheral nervous system, or specific to any other organ (other), with heart a cardiovascular system terms as well as brain and peripheral nervous system terms being grouped for simplicity.

### 1.1.6 Digenic Pairs within 11 CHD-Associated Genes

Expected number of digenic pairs of PTCs was calculated empirically based on the per gene probability of having a PTC; P(gene_A) = PTC_Freq_cases(gene_A)/n_cases. We then assumed the probability of having a PTC in each individual gene is independent, such that the probability of having any cross-gene pair of variants is equal to the probability of having a PTC in one gene times the probability of having a PTC in another gene - P(gene.A & gene.B) = P(gene_A) * P(gene_B). Finally, we summed the probabilities across gene pairs without replacement and without considering order; probability of having digenic pair across CHD associated genes I through J = P(PTCpair) = Sum(I to J)[P(gene_A) * P(gene_B)].

## Supporting information

AdditionalFile1

AdditionalFile2

AdditionalFile3

AdditionalFile4

AdditionalFile5

## Data Availability

All data produced in the present study are available upon reasonable request to the authors or available online from dbGaP

## Bibliography

1. Triedman, J. K. & Newburger, J. W. Trends in congenital heart disease: The next decade. Circulation 133, 2716–2733 (2016).

2. Botto, L. D. et al. Seeking causes: Classifying and evaluating congenital heart defects in etiologic studies. Birth Defects Res. A Clin. Mol. Teratol. 79, 714–727 (2007).

3. Ferencz, C. et al. Congenital heart disease: Prevalence at livebirth. Am. J. Epidemiol. 121, 31–36 (1985).

4. Hoffman, J. I. E. & Kaplan, S. The incidence of congenital heart disease. J. Am. Coll. Cardiol. 39, 1890–1900 (2002).

5. Marelli, A. J., Mackie, A. S., Ionescu-Ittu, R., Rahme, E. & Pilote, L. Congenital heart disease in the general population: changing prevalence and age distribution. Circulation 115, 163–172 (2007).

6. Zhu, W. & Lo, C. W. Insights into the genetic architecture of congenital heart disease from animal modeling. Zool. Res. 44, 577–590 (2023).

7. Zaidi, S. & Brueckner, M. Genetics and genomics of congenital heart disease. Circ. Res. 120, 923– 940 (2017).

8. Mani, A. Pathogenicity of DE Novo rare variants: Challenges and opportunities. Circ. Cardiovasc. Genet. 10, (2017).

9. MacArthur, D. G. et al. A systematic survey of loss-of-function variants in human protein-coding genes. Science 335, 823–828 (2012).

10. Miller, J. N. & Pearce, D. A. Nonsense-mediated decay in genetic disease: friend or foe? Mutat. Res. Rev. Mutat. Res. 762, 52–64 (2014).

11. Backwell, L. & Marsh, J. A. Diverse molecular mechanisms underlying pathogenic protein mutations: Beyond the loss-of-function paradigm. Annu. Rev. Genomics Hum. Genet. 23, 475–498 (2022).

12. Khajavi, M., Inoue, K. & Lupski, J. R. Nonsense-mediated mRNA decay modulates clinical outcome of genetic disease. Eur. J. Hum. Genet. 14, 1074–1081 (2006).

13. Klonowski, J., Liang, Q., Coban-Akdemir, Z., Lo, C. & Kostka, D. Aenmd: Annotating escape from nonsense-mediated decay for transcripts with protein-truncating variants. Bioinformatics 39, (2023).

14. Karczewski, K. J. et al. Author Correction: The mutational constraint spectrum quantified from variation in 141,456 humans. Nature 590, E53 (2021).

15. Cummings, B. B. et al. Transcript expression-aware annotation improves rare variant interpretation. Nature 581, 452–458 (2020).

16. Aoki, Y. et al. Gain-of-function mutations in RIT1 cause Noonan syndrome, a RAS/MAPK pathway syndrome. Am. J. Hum. Genet. 93, 173–180 (2013).

17. Tartaglia, M. et al. PTPN11 mutations in Noonan syndrome: molecular spectrum, genotype-phenotype correlation, and phenotypic heterogeneity. Am. J. Hum. Genet. 70, 1555–1563 (2002).

18. Wu, M. C. et al. Rare-variant association testing for sequencing data with the sequence kernel association test. Am. J. Hum. Genet. 89, 82–93 (2011).

19. Homsy, J. et al. De novo mutations in congenital heart disease with neurodevelopmental and other congenital anomalies. Science 350, 1262–1266 (2015).

20. Jin, S. C. et al. Contribution of rare inherited and de novo variants in 2,871 congenital heart disease probands. Nat Genet 49, 1593–1601 (2017).

21. Morton, S. U. et al. Association of Potentially Damaging De Novo Gene Variants With Neurologic Outcomes in Congenital Heart Disease. JAMA Netw Open 6, e2253191 (2023).

22. Wang, Y. J. et al. Systems analysis of de novo mutations in congenital heart diseases identified a protein network in the hypoplastic left heart syndrome. Cell Syst. 13, 895–910.e4 (2022).

23. Ma, L., Lu, M.-F., Schwartz, R. J. & Martin, J. F. Bmp2 is essential for cardiac cushion epithelial-mesenchymal transition and myocardial patterning. Development 132, 5601–5611 (2005).

24. Rivera-Feliciano, J. & Tabin, C. J. Bmp2 instructs cardiac progenitors to form the heart-valve-inducing field. Dev Biol 295, 580–588 (2006).

25. Singh, R. et al. Tbx2 and Tbx3 induce atrioventricular myocardial development and endocardial cushion formation. Cell Mol Life Sci 69, 1377–1389 (2012).

26. Saxon, J. G. et al. BMP2 expression in the endocardial lineage is required for AV endocardial cushion maturation and remodeling. Dev Biol 430, 113–128 (2017).

27. Durst, R. et al. Mutations in DCHS1 cause mitral valve prolapse. Nature 525, 109–113 (2015).

28. McBride, K. L. et al. NOTCH1 mutations in individuals with left ventricular outflow tract malformations reduce ligand-induced signaling. Hum Mol Genet 17, 2886–2893 (2008).

29. Stanley, K. J. et al. Expanding the phenotypic spectrum of NOTCH1 variants: clinical manifestations in families with congenital heart disease. Eur J Hum Genet 32, 795–803 (2024).

30. Yang, A. et al. CHDgene: A curated database for congenital heart disease genes. Circ. Genom. Precis. Med. 15, e003539 (2022).

31. Osborne, L. R. & Mervis, C. B. Rearrangements of the Williams-Beuren syndrome locus: molecular basis and implications for speech and language development. Expert Rev Mol Med 9, 1–16 (2007).

32. Rots, D. et al. The clinical and molecular spectrum of the KDM6B-related neurodevelopmental disorder. Am J Hum Genet 110, 963–978 (2023).

33. Szot, J. O. et al. A screening approach to identify clinically actionable variants causing congenital heart disease in exome data. Circ. Genom. Precis. Med. 11, e001978 (2018).

34. Fregeau, B. et al. De Novo mutations of RERE cause a genetic syndrome with features that overlap those associated with proximal 1p36 deletions. Am. J. Hum. Genet. 98, 963–970 (2016).

35. Jordan, V. K. et al. Genotype-phenotype correlations in individuals with pathogenic RERE variants. Hum. Mutat. 39, 666–675 (2018).

36. Kim, B. J. et al. An allelic series of mice reveals a role for RERE in the development of multiple organs affected in chromosome 1p36 deletions. PLoS One 8, e57460 (2013).

37. Malgapo, M. I. P. & Linder, M. E. Substrate recruitment by zDHHC protein acyltransferases. Open Biol 11, 210026 (2021).

38. Bell, C., Nevitt, S., McKay, V. H. & Fattah, A. Y. Will the real Moebius syndrome please stand up? A systematic review of the literature and statistical cluster analysis of clinical features. Am J Med Genet A 179, 257–265 (2019).

39. Liu, J. et al. A dual role for ErbB2 signaling in cardiac trabeculation. Development 137, 3867–3875 (2010).

40. Sanchez-Soria, P. & Camenisch, T. D. ErbB signaling in cardiac development and disease. Semin Cell Dev Biol 21, 929–935 (2010).

41. Zhang, J.-R., Li, X.-X., Hu, W.-N. & Li, C.-Y. Emerging Role of TRIM Family Proteins in Cardiovascular Disease. Cardiology 145, 390–400 (2020).

42. Lin, L., Bu, L., Cai, C.-L., Zhang, X. & Evans, S. Isl1 is upstream of sonic hedgehog in a pathway required for cardiac morphogenesis. Dev Biol 295, 756–763 (2006).

43. Meyers, E. N. & Martin, G. R. Differences in left-right axis pathways in mouse and chick: functions of FGF8 and SHH. Science 285, 403–406 (1999).

44. Wiegering, A., Rüther, U. & Gerhardt, C. The Role of Hedgehog Signalling in the Formation of the Ventricular Septum. J Dev Biol 5, (2017).

45. Washington Smoak, I., et al. Sonic hedgehog is required for cardiac outflow tract and neural crest cell development. Dev Biol 283, 357–372 (2005).

46. Li, Y. et al. Global genetic analysis in mice unveils central role for cilia in congenital heart disease. Nature 521, 520–524 (2015).

47. The ERBB2 c.1795C>T, p.Arg599Cys variant is associated with left ventricular outflow tract obstruction defects in humans. Human Genetics and Genomics Advances 6, 100446 (2025).

48. Leung, C. et al. Rac1 Signaling Is Required for Anterior Second Heart Field Cellular Organization and Cardiac Outflow Tract Development. J Am Heart Assoc 5, (2015).

49. Tavares, A. L. P., Mercado-Pimentel, M. E., Runyan, R. B. & Kitten, G. T. TGF beta-mediated RhoA expression is necessary for epithelial-mesenchymal transition in the embryonic chick heart. Dev Dyn 235, 1589–1598 (2006).

50. Hamanaka, K. et al. De Novo Truncating Variants in the Last Exon of SEMA6B Cause Progressive Myoclonic Epilepsy. Am J Hum Genet 106, 549–558 (2020).

51. Kim, H. J. et al. Heterozygous frameshift variants in HNRNPA2B1 cause early-onset oculopharyngeal muscular dystrophy. Nat Commun 13, 2306 (2022).

52. Stout, K. K. et al. 2018 AHA/ACC Guideline for the Management of Adults With Congenital Heart Disease: Executive Summary: A Report of the American College of Cardiology/American Heart Association Task Force on Clinical Practice Guidelines. J Am Coll Cardiol 73, 1494–1563 (2019).

53. Kousi, M. et al. Evidence for secondary-variant genetic burden and non-random distribution across biological modules in a recessive ciliopathy. Nat Genet 52, 1145–1150 (2020).

54. Gabriel, G. C., Young, C. B. & Lo, C. W. Role of cilia in the pathogenesis of congenital heart disease. Semin Cell Dev Biol 110, 2–10 (2021).

55. Sullivan, J. M., De Rubeis, S. & Schaefer, A. Convergence of spectrums: neuronal gene network states in autism spectrum disorder. Curr Opin Neurobiol 59, 102–111 (2019).

56. Liu, X. et al. The complex genetics of hypoplastic left heart syndrome. Nat Genet 49, 1152–1159 (2017).

57. Yoshida, Y. et al. A genetic and developmental biological approach for a family with complex congenital heart diseases-evidence of digenic inheritance. Front Cardiovasc Med 10, 1135141 (2023).

58. Pittman, M. E. Computational approaches identify novel risk loci and interactions in heart defects. (UCSF, 2023).

59. Gifford, C. A. et al. Oligogenic inheritance of a human heart disease involving a genetic modifier. Science 364, 865–870 (2019).

60. Akhirome, E., et al. The Genetic Architecture of a Congenital Heart Defect Is Related to Its Fitness Cost. Genes (Basel) 12, (2021).

61. Kim, B. J. et al. RERE deficiency leads to decreased expression of GATA4 and the development of ventricular septal defects. Dis Model Mech 11, (2018).

