## AdditionalFile4 for "Analysis of Premature Termination Codons Predicted to Escape Nonsense Mediated Decay Identifies Novel Genes, Pathways, and Networks Contributing to an Oligogenic Etiology of Congenital Heart Disease"

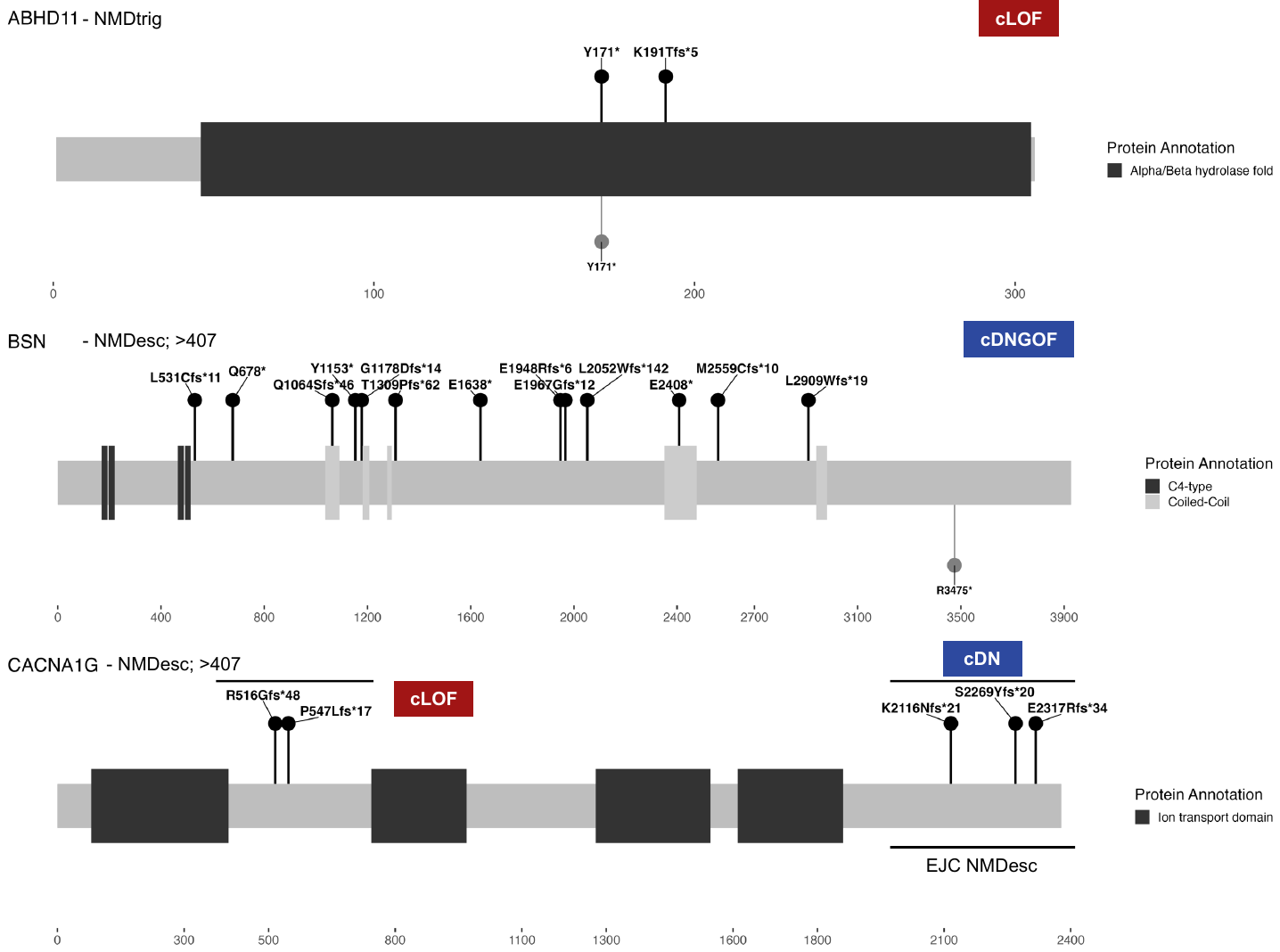

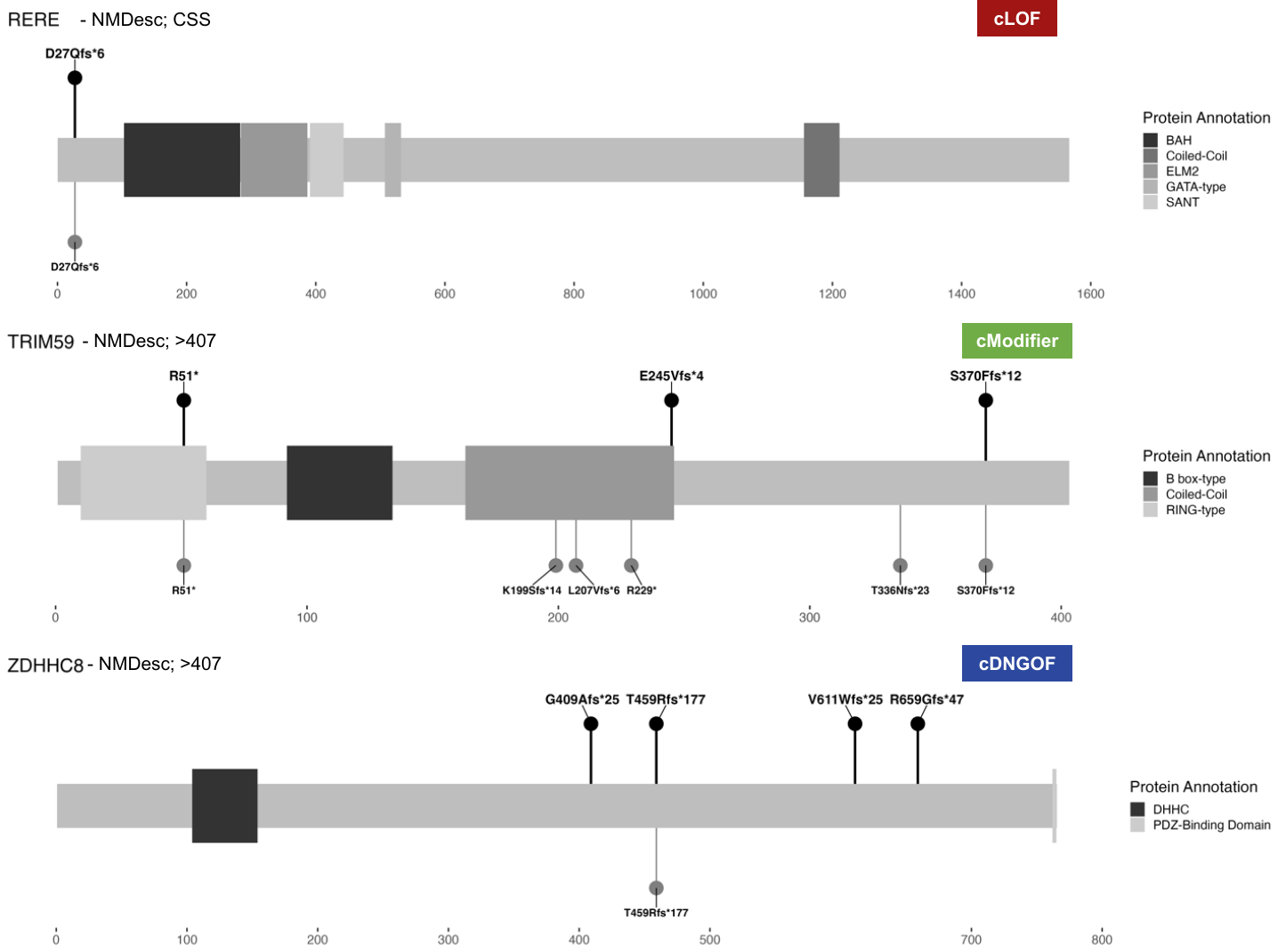

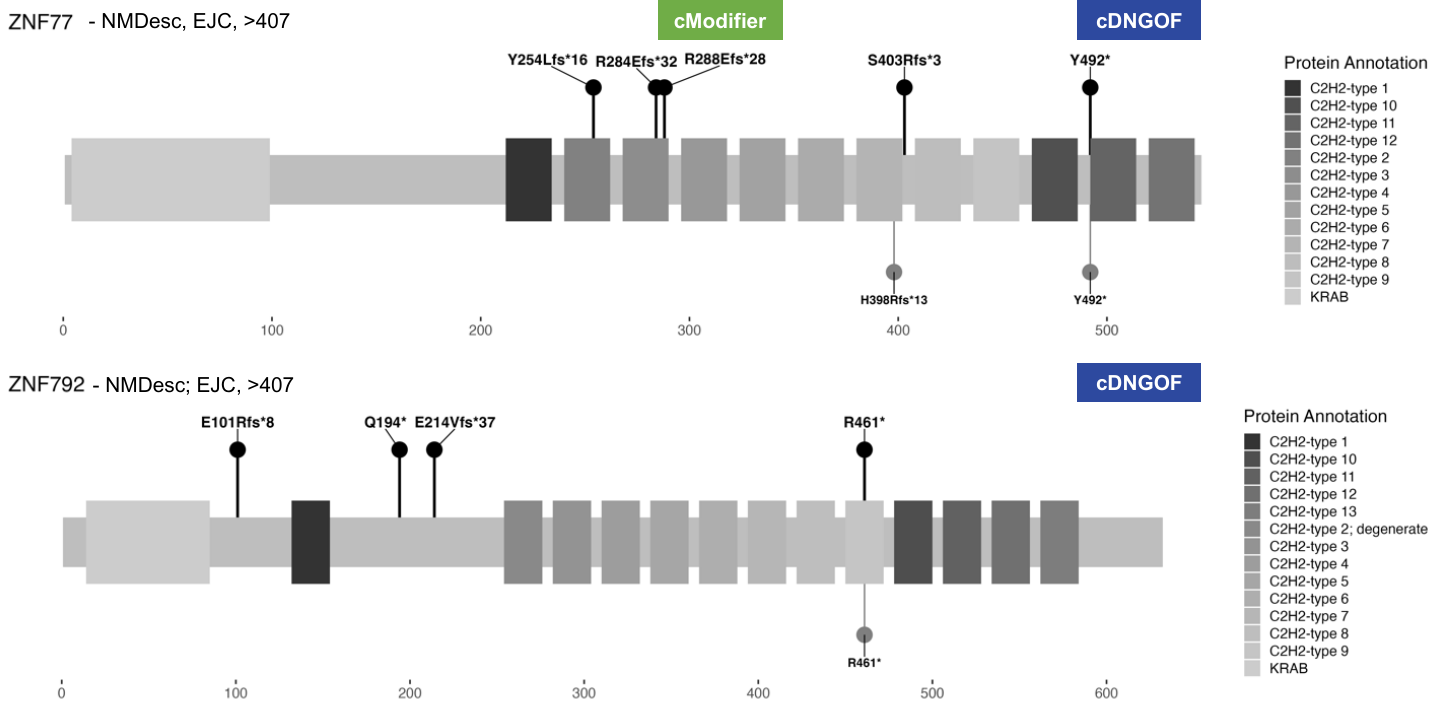
 **Supplimental Figure 1.** **Eleven CHD-Associated Genes Recovered from NMD PTC Analysis and Their Speculated Molecular Effect.** Cartoon representation of the protein encoded by our 11 CHD assocaited genes, the PTC variants found and their speculated molecular effect. For the 3 genes recovered from NMDtrig analysis (ABHD11, FLT4, KDM6B), these were assigned as candidate Loss of Function (cLOF). For predicating molecular effects of the NMDesc recovered genes, NMDesc was assumed, leading to most receiving the candidate Dominant Negative or Gain of Function (cDNGOF) classification; however, some genes were not given this label. For **CACNA1G**, the first 2 PTCs would remove ion channel domains required for a viable protein and were therefore labeled as cLOF. The remaining PTCs fall in the final exon of the transcript (NMDesc by >407 and EJC) and would not cut out the ion channel domains, instead affecting N-terminus regulatory sequences. Further, no distinct phenotypic differences were recognized between the patients with PTCs in these two reigns. Therefore, the candidate dominant negative (cDN) label for these final exon localized PTCs. Although protein produced by **MATN4**’s PTC-inflicted transcripts do not appear to be able to create a meaning amino acid sequence, further analysis suggest these PTCs may result in cDNGOF. In this case, if the CSS rule would be relaxed from the first 100bp of the transcript to the first 225bp (a reasonable number evident from previous studies) there are downstream inframe coding start site that the translation machinery could begin translating from (sequence from CCDS13348), potentially leading to a c-terminally truncated protein. This is in contrast to **RERE**, whose PTCs are NMDesc under the CSS rule but whose transcripts contain many out of frame coding start sites downstream of the observed PTCs before there is an inframe coding start site. This would vastly reduce the likelihood of a native, but C-terminal truncated protein to be produced, earning **RERE** the cLOF label. We could not come up with a reasonable prediction for **TRIM59**, given that there was no correlation between the location (more c-terminal vs more n-terminal) or the PTCs and the patient phenotype. Further, while statistically significant most of the PTCs are also seen in controls (both in cohort and gnomAD), thus we predict that **TRIM59** likely contributes to the genetic architecture of CHD without driving CHD (cModifier). For **ZNF77**, while each individual PTC found was ultra rare, PTCs in EJC region are overall pretty common. In the case of **ZNF77**, PTCs in this region have been associated with Meckel-Bruber syndrome, coronary heart disease, height and growth, fibromyalgia syndrome, and other phenotypes, as well as clinically relevant biologic impacts (10.1007/s43032-021-00835-5, 10.1371/journal.pone.0128348, 10.1038/s41467-018-06148-7, 10.3390/diagnostics12102561, 10.1371/journal.pone.0065033). As such, we have them the cModifier and cDNGOF labels due to this relatively common appearance of PTCs paired with evidence implicating them in disease. Finally, BSN and ZNF792, we could not learn much from the distribution of PTCs and as such, these were given the cDNGOF label under the assumption that truncated protein would be expressed.

**Key**: Each lollipop represents a unique PTC-induced amino acid change, with those found in cases being displayed on the positive y-axis and controls on the negative y-axis. Each lollipop has the corresponding HGVS protein annotation (xPOSyEF*DIS ; x - amino acid symbol, POS - amino acid position, y - amino acid change, EF - effect of variant with fs for frameshift, * - denoting a stop codon, DIS - distance to the generated stop codon).

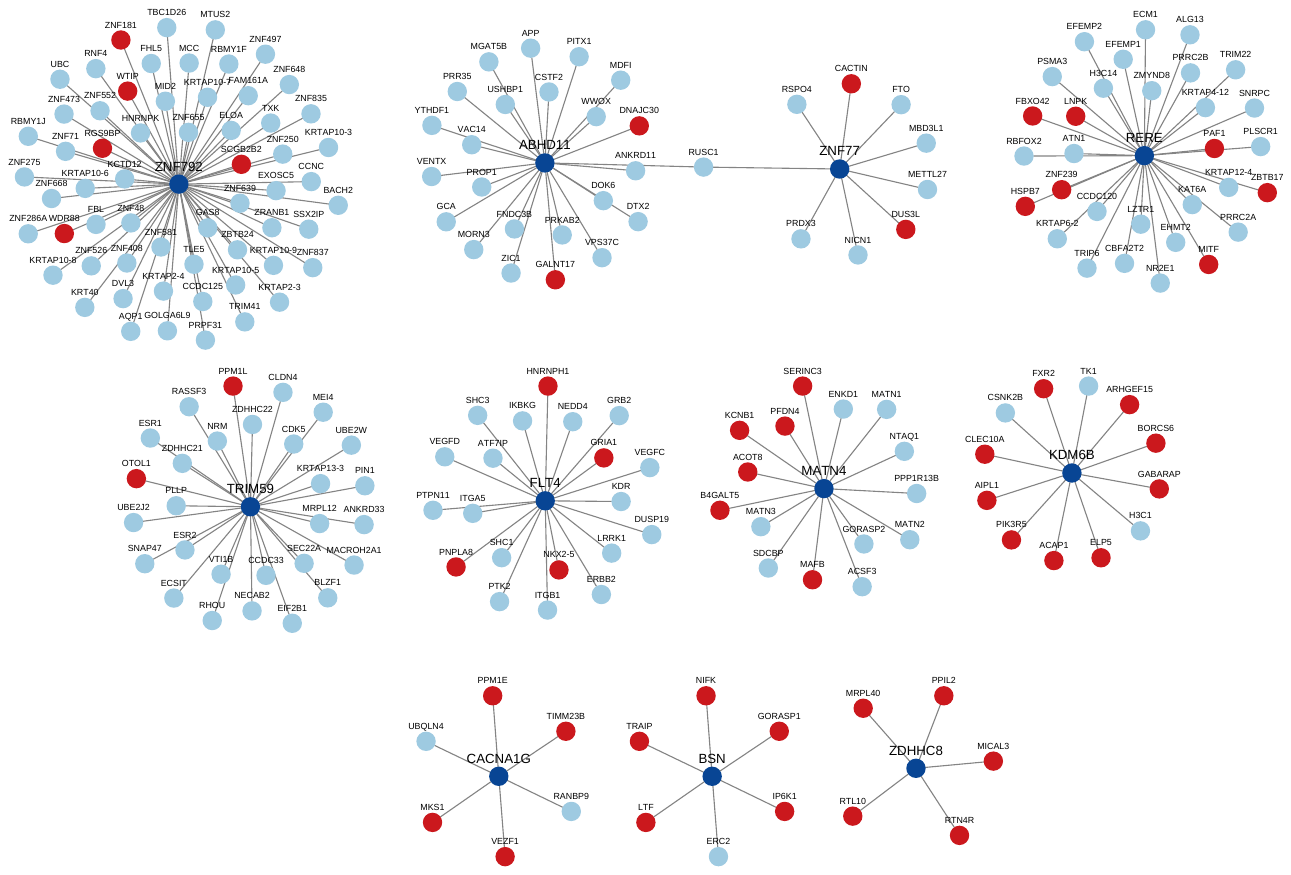

**Supplemental Figure 2. Protein Interaction Network for all 11 CHD-Associated Genes and Their Primary Interactors.** Protein-protein interaction network of our 11 CHD genes and their primary known or predicted protein interactors. Dark blue central nodes are our 11 CHD genes recovered from burden analysis of NMDtrig or NDMesc PTCs. Light blue nodes are primary known interactors. Red nodes are predicted interactors.

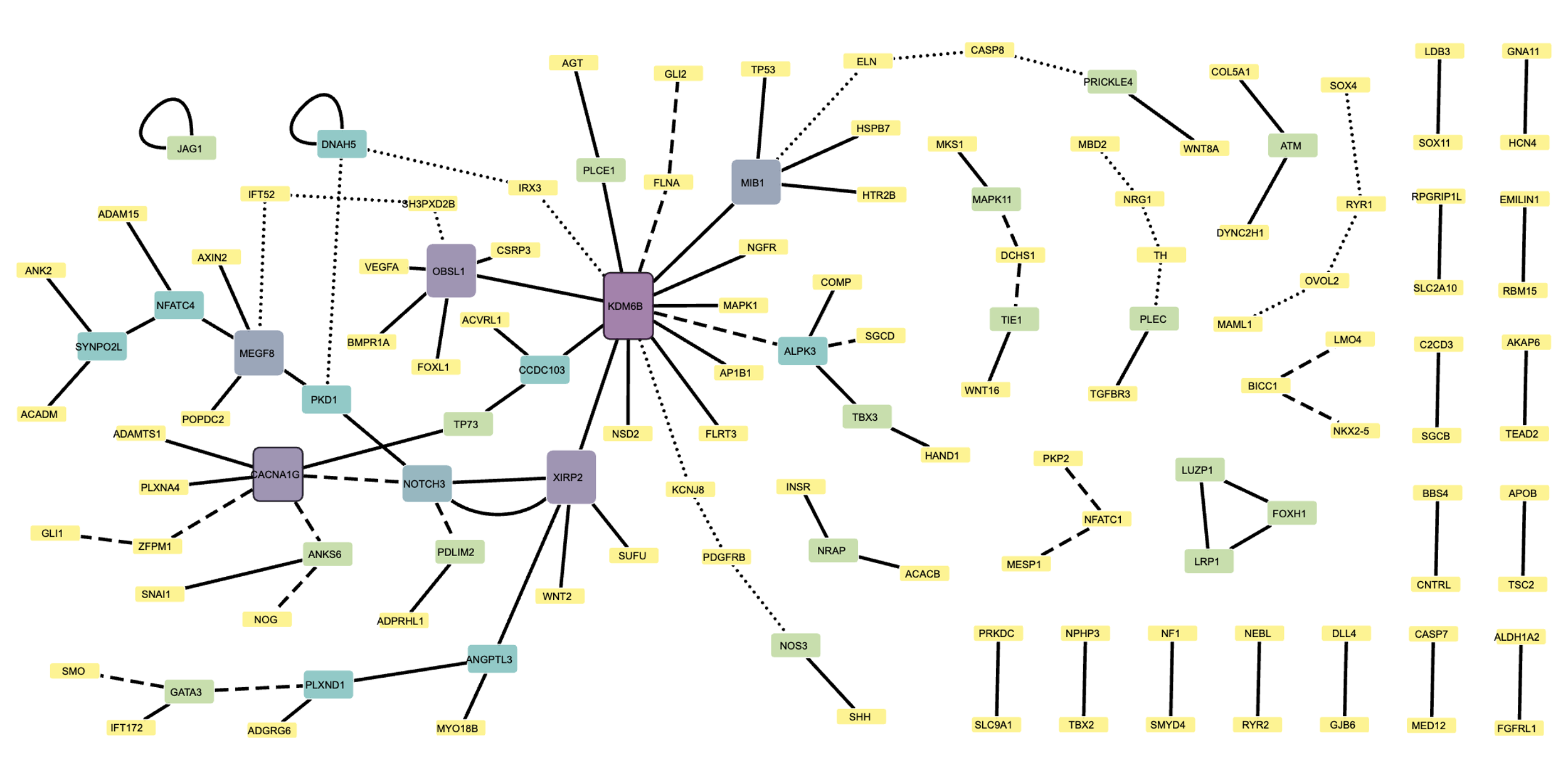

**Supplemental Figure 3. All Oligogenic Combinations of PTCs in Heart Development Genes found in CHD patients.** Heart development associated gene-sets previously recovered from PTC-NMD analysis were further interrogated and found to have increased incidence of oligogenic occurrence, observing digenic, trigenic and quad-genic combinations. Each combination was unique combination except for 2 patients with a *NOTCH3*-*XIRP2* PTC pair. Notable is KDM6B, seen in 14 unique oligogenic sets corresponding to 14 patients.
